# Use of Computerized Neurocognitive Assessment Software for the Detection of Alcohol Intoxication

**DOI:** 10.1101/2020.05.12.20086868

**Authors:** Sanjeev Janarthanan, Huy Phi, Benjamin Flores, Yael Katz, David M. Eagleman, Bin Huang, Reza Hosseini Ghomi

## Abstract

**Background:** Acute ingestion of alcohol impairs cognitive function and poses significant threat to public health and safety with impaired operation of motor vehicles. However, there is a lack of access to tools to assess one’s cognitive impairment due to alcohol. The purpose of this study was to explore the use of a neuropsychological assessment software, BrainCheck, to assess levels of alcohol impairment based on performance on the neuropsychological assessments.

**Methods:** The BrainCheck battery was administered to 91 volunteer participants. Participants were required to take a baseline battery prior to any alcohol ingestion, and another testing battery after a voluntary drinking period. Blood alcohol content (BAC) for the participants was obtained using a breathalyzer. Participant assessments were grouped into an intoxicated group with BAC > 0.05, and a sober group with BAC ≤ 0.05. Statistical analysis comparing intoxicated vs sober performance on the BrainCheck battery was performed. Significant metrics deemed by relative t-test and correlations with BAC were used to build statistical learning models to predict participant BAC and classify intoxicated vs. sober.

**Results:** Relative t-tests found two assessment metrics to be significant among comparison groups after P-value correction. Four test metrics were observed to moderately correlate (|*r*| > 0.40) with BAC levels. Three linear regression models (least-squares, ridge and LASSO) were built to predict participant BAC levels, with the best performing model being the least-squares model with a RMSE of 0.027, compared to the baseline RMSE of 0.061. Logistic regression models were built to classify whether the participant was intoxicated or not, with the best model performing at 80.6% accuracy, 73.3% sensitivity, and 75.0% specificity.

**Discussion:** The BrainCheck battery has potential to predict alcohol impairment, including participant BAC levels and if the participant is intoxicated or not. BrainCheck provides another option to assess an individual’s cognitive impairment due to alcohol, with the utility of being portable and available on one’s smartphone.

## Background

Alcohol is a widely known central nervous system depressant, and acute consumption of alcohol results in impaired cognitive and psychomotor function, including reduced attention, alterations in memory, and reaction time (Valenzuela, 1997). The acute effects of alcohol pose significant threats to public health and safety. In 2018, 10,511 people died in alcohol-related motor vehicle accidents, accounting for 29% of all traffic-related deaths in the United States (NHTSA, 2019). Although most legal limits for driving are set at 0.08 % blood alcohol content (BAC), research has shown that human subjects with BAC of 0.05 % display significantly diminished performance on psychomotor tasks of attention and reaction time compared to controls (*p* < 0.05) and even greater significance for BAC of 0.08% (*p* < 0.01) (Grant et al., 2000).

Conventional methods to detect alcohol impairment include the Standardized Field Sobriety Test which typically requires a police officer to administer the test at the roadside (Stuster, 1998), however this method does not prevent drivers from operating their vehicles. Breathalyzers can also be used to calculate an individual’s BAC, and although personal breathalyzers are becoming increasingly more accessible, they are still expensive, require calibration every 6-12 months, and lack consistency compared to police-grade breathalyzers (Riordan et al., 2017). Thus, a highly sensitive, rapid and self-administered cognitive screening test could aid in early detection of alcohol impairment, and prevent an intoxicated driver from operating a motor vehicle. Cognitive deficits associated with a BAC of 0.05% have been shown to be a reference for cognitive deficits associated with mild traumatic brain injury (mTBI) on neuropsychological assessment performance (Collie et al., 2002). Prior work on computerized neuropsychological assessment batteries have also been able to assess impairment in cognitive functions with acute alcohol consumption (Schweizer et al., 2006), however this study was only used to assess performance on the neuropsychological assessment, and not in predicting the users level of alcohol impairment.

We explore the use of BrainCheck, a computerized neurocognitive assessment software that is available on iPad, iPhone or desktop browser. The BrainCheck testing battery administers neurocognitive tests, which work to maximize diagnostic accuracy, portability and ease of operator use. BrainCheck Sport has previously been validated for diagnostic accuracy for detection of mTBI, and BrainCheck Memory in identifying cognitive impairment and dementia (Yang et al., 2017; Groppell et al., 2019). Our aim is to test the utility of the BrainCheck battery on detecting acute alcohol impairment by comparing BrainCheck assessment composite scores with BAC obtained from participant breathalyzer scores.

## Methods

### Assessment selection

The BrainCheck battery contains five assessments, described in Table 1. Assessments are derived from traditional neuropsychological tests, including the Flanker test, Digit Symbol Substitution, Stroop, and the Trail Making A & B tests (Eriksen & Eriksen, 1974; Jaeger, 2018; Scarpina & Tagini, 2017; Bowie & Harvey, 2006). The coordination test is adapted from the Balance Error Scoring System (Bell et al., 2011).

**Table 1.**
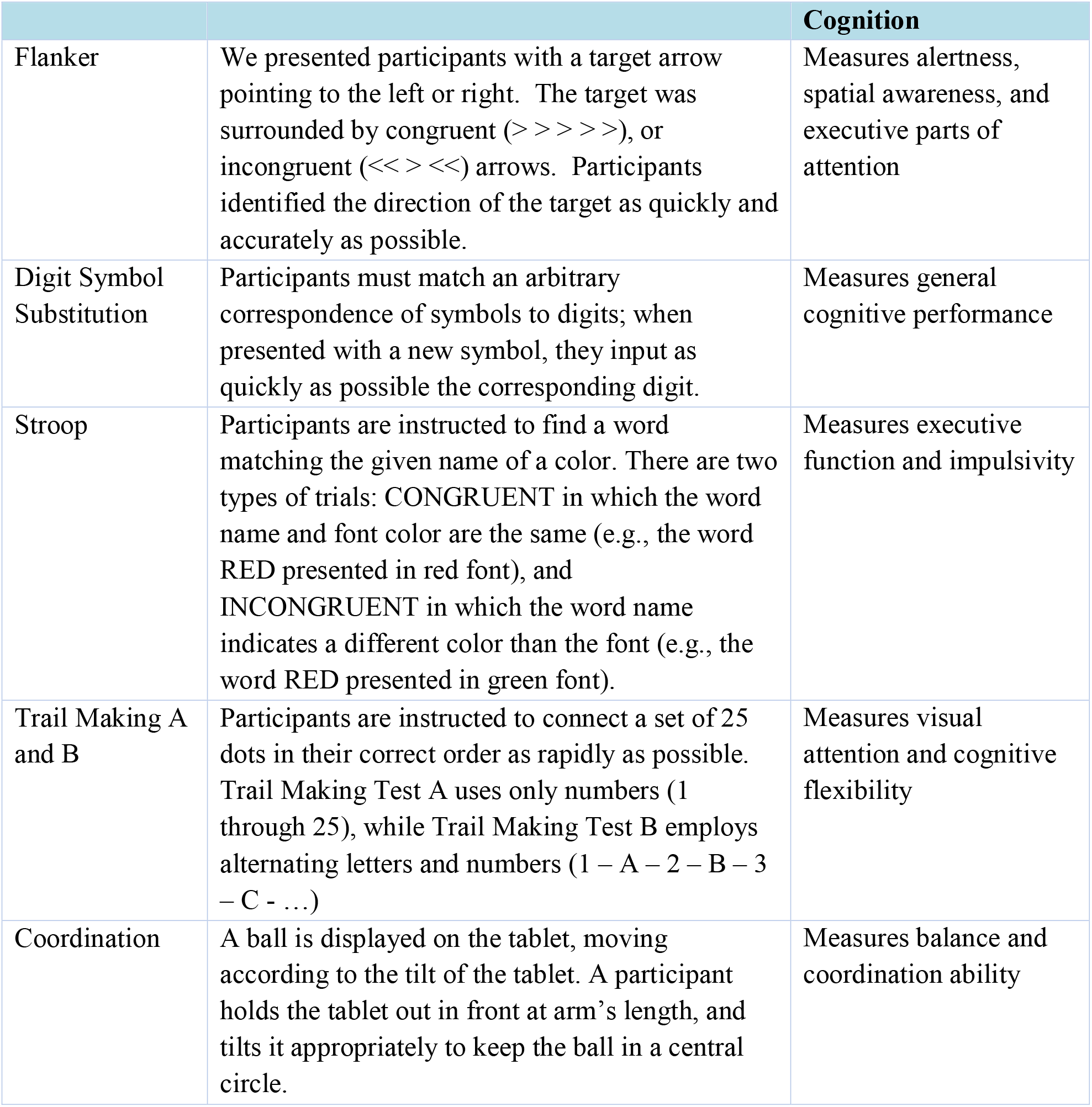
Neuropsychological tests in the BrainCheck battery

### Inclusion criteria

All participants required the use of both arms and legs, and perfect/corrected vision. Participants needed to be of legal drinking age (21), and have a breathalyzer BAC of 0.00 immediately prior to the first set of assessments.

Excluded participants were individuals with prior exposure to the battery, or those who admitted to alcohol or drug use within the previous 6 hours. Further exclusion criteria were individuals with impaired function of upper/lower extremities, memory disorders, imperfect/uncorrected vision or those who received less than 4 hours of sleep the previous night.

### Obtaining participants

To obtain participants, likely candidates were approached in public spaces and asked if they would volunteer for the study. Volunteers were collected through interoffice relationships, and random participants selected from a local pub in downtown Houston, Texas. Participants were required to take a baseline BrainCheck battery prior to any alcohol consumption, then took the battery a second time after an alcohol ingestion period of approximately 5-6 hours. Participants were not instructed or encouraged to consume alcohol. Only those who were consuming alcohol of their own volition regardless of their involvement in the study were asked to participate.

#### Statistical Analysis

Relative sample t-tests were performed to compare assessment performance among participants for Test 1 before alcohol consumption and Test 2 after the alcohol consumption period. The Sidak correction method of t-tests for multiple comparisons was used to correct and define a new significance value for α (Blakesley et al., 2009). Additionally, we calculated the difference of each assessment metric for each participant before and after alcohol consumption, and calculated the Pearson correlation coefficient (*r*) between them and the participant’s BAC after consuming alcohol.

Participant assessments were placed into two groups, those with a BAC above 0.05% comprised the intoxicated group, and those below or equal that threshold were in the sober groups. For each assessment, we again used relative sample t-tests and Sidak correction method to compare the assessment metrics of the intoxicated group with the sober group.

Linear regression models were built, including least-squares regression, ridge regression, and least absolute shrinkage selection operator (LASSO) regression, to predict BAC based on the difference of assessment metrics before and after consuming alcohol. To minimize the impact of poor features and prevent overfitting, we applied L1 (Ridge) and L2 (LASSO) penalties to the models to restrict the feature weights (Hastie et al., 2001). For the Ridge regression model, we tuned the L2 penalty from 0.0001 to 100,000, while we turned the L1 penalty from 0.0000001 to 10 in the LASSO regression model. We split 70% of the data into the training set, 10% into the validation set, and 20% into the test set. We used the validation data to evaluate the best L1 and L2 penalties for our LASSO and ridge models respectively.

Logistic Regression models with regularizations (L1, L2 and elastic net) were built to classify participants into the sober and intoxicated groups. We split 80% of the data into the training set and the other 20% into the testing set, based on the unique tester ID assigned to each participant. We evaluated the performance of the model using the receiver operator characteristic (ROC) curve and located the optimal threshold for the logistic model with maximum sensitivity/specificity.

Data analysis was performed using the Python programming language.

### Results

#### Demographics

We recruited 91 participants. This sample ended up being mostly individuals within the age range of 21-70 years old (47.3 % female). Participant demographics are displayed in Table 2. Mean BAC among all participants after the drinking period was 0.0997 (SD = 0.0373).

**Table 2.** Participant Demographics

| | Sex<br>(Male/Female) | Age<br>(Mean ± SD) | BAC after Drinking<br>Period (Mean ± SD) |
| --- | --- | --- | --- |
| Participants (n = 91) | 48/43 | 34.37 ± 9.41 | 0.0997 ± 0.0373 |

#### Assessment Metrics

All assessment metrics are displayed in Table 3. The Sidak method correction for multiple t-tests defined a new threshold for significant p-values, at α = 0.005. We found that metrics that are significantly different (*p* < 0.005) before and after alcohol consumption are the mean of Trails A duration and median of Trails A duration. Metrics that are significantly different for the intoxicated versus sober groups (*p* < 0.005) are the same. Boxplots that display the separation for metrics with a *p* < 0.1 are shown in Figure 1. We found four metrics (median of digit symbol duration, mean of digit symbols correct per second, mean of stroop reaction time and the median of incongruent stroop reaction time) with |*r|* > 0.40 (Figure 2), which would be considered as moderate correlations with BAC (Schober et al., 2018).

**Table 3.** Comparison of assessment metrics. *P*-values are from relative t-tests between sober vs intoxicated groups, intoxicated being BAC > 0.05, *p*-values comparing performance of participants before and after the drinking period, and *r* values are correlations between the change in the metric measurement and BAC levels. Bolded values are for *p*-values < 0.005 according to Sidak corrected *α* value, and |*r*| > 0.40.

| Assessment Metric | $P$-value for comparing Intoxicated vs. Sober groups | $P$-value for comparing Test 1 vs. Test 2 groups | Pearson Correlation Coefficient ($r$) | Description |
| --- | --- | --- | --- | --- |
| Median of Digit Symbol Duration | 0.16 | 0.19 | **-0.45** | Total time taken to complete assessment. |
| Mean of Digit Symbols Correct per Second | 0.20 | 0.26 | **0.41** | Mean of correct responses in a second |
| Mean of Trails A Duration | **0.004** | **0.002** | -0.31 | Average of response times between each click for Trail Making A |
| Mean of Trails B Duration | 0.19 | 0.17 | -0.36 | Average of response times between each click for Trail Making B |
| Median of Trails A Duration | **0.0027** | **0.00005** | -0.10 | Median of response times between each click for Trail Making A |
| Median of Trails B Duration | 0.16 | 0.05 | -0.25 | Median of response times between each click for Trail Making B |
| Mean of Stroop Reaction Time | 0.083 | 0.06 | **-0.47** | Average reaction time for all responses |
| Median of Incongruent Stroop Reaction Time | 0.096 | 0.04 | **-0.44** | Median reaction time for all responses |
| Mean of Correct Flanker Reaction Time | 0.072 | 0.08 | -0.040 | Median reaction time of all correct responses |
| Median of Correct Flanker Reaction Time | 0.094 | 0.10 | -0.17 | Median reaction time of all responses when arrows point in the same direction |
| Mean of Balance Distance from Center | 0.30 | 0.23 | -0.22 | Mean radial distance from center circle |

**Figure 1.**
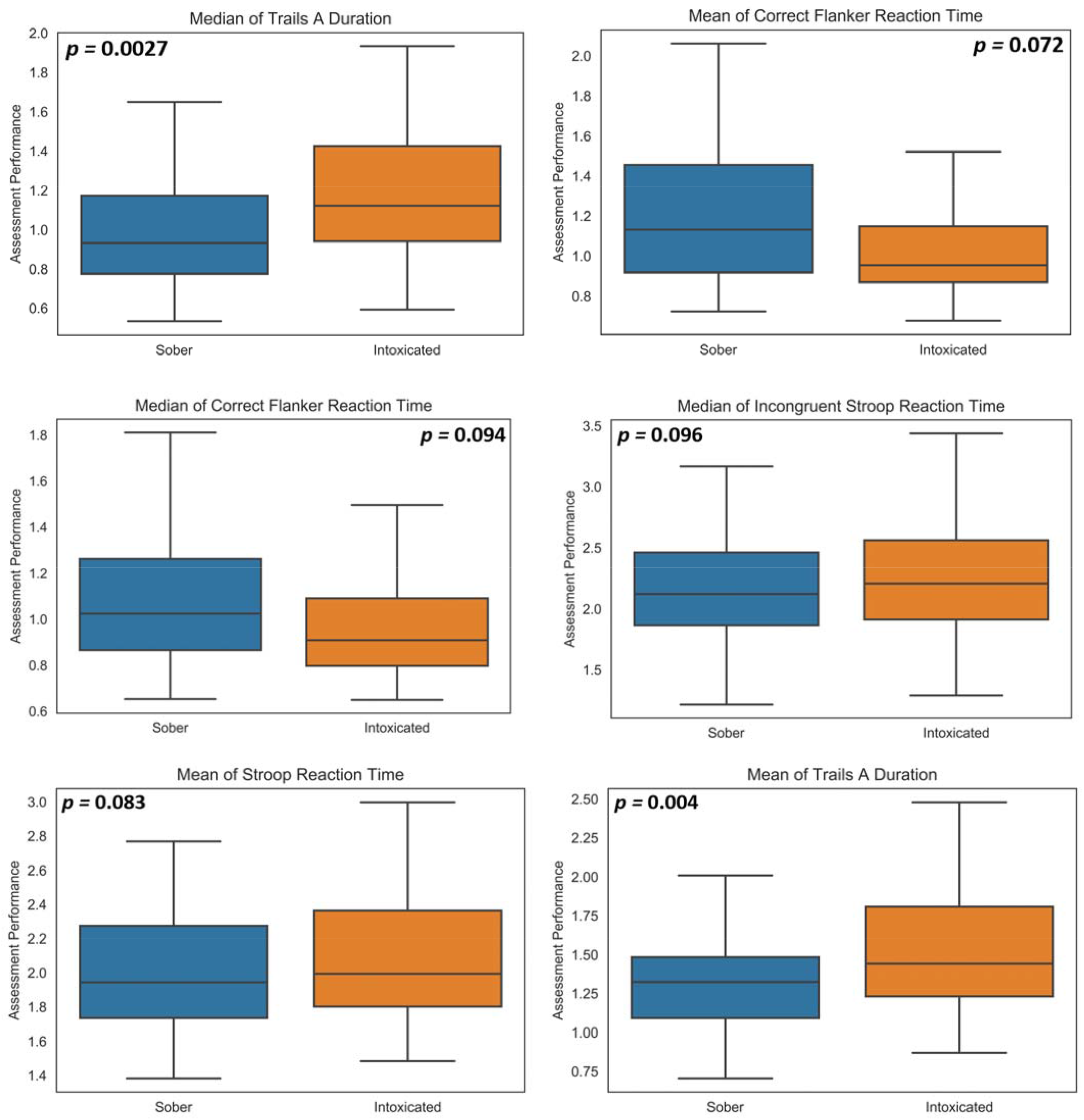
Boxplot of the comparison assessment metrics between intoxicated and sober groups with *p*-values < 0.1

**Figure 2.**
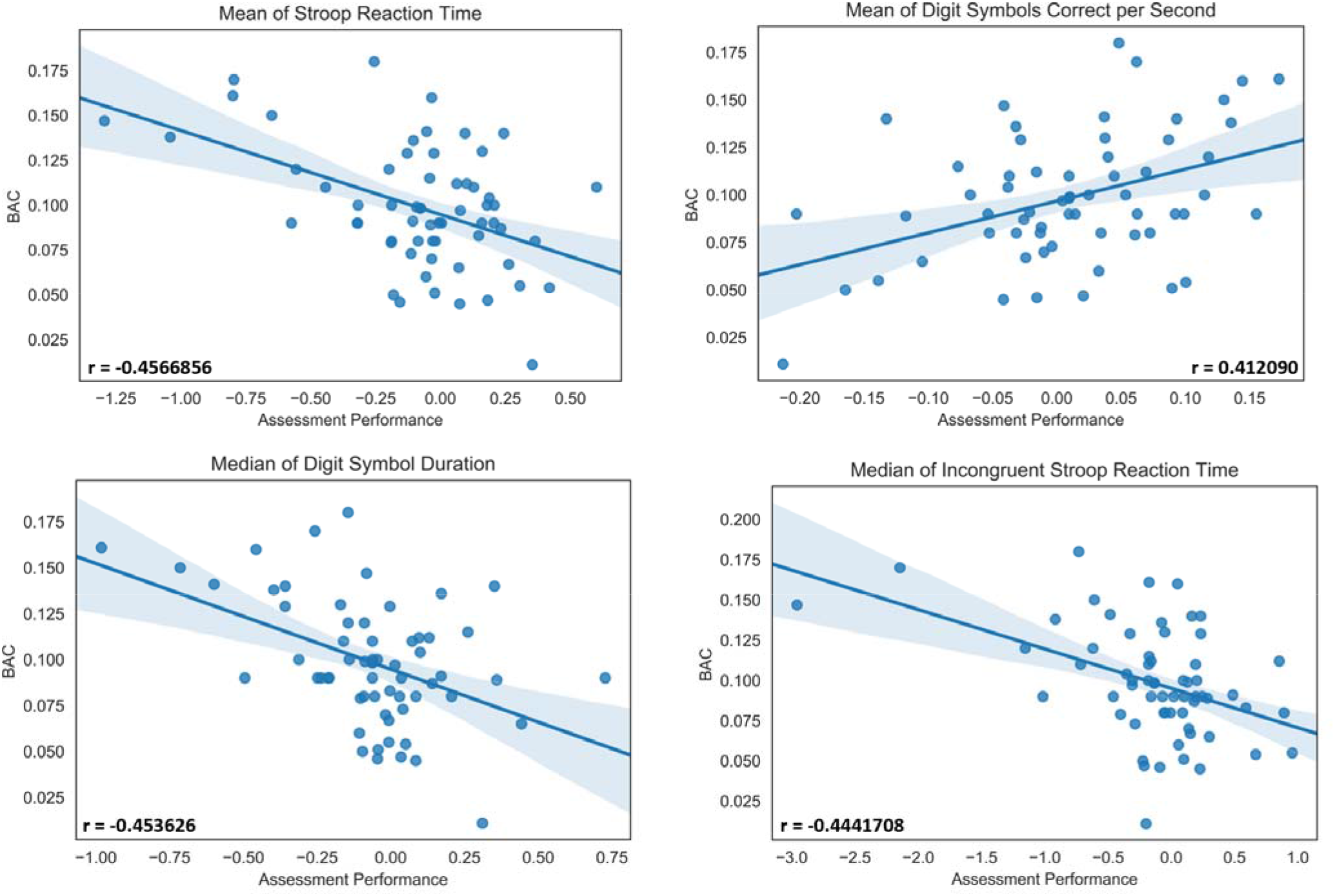
Correlations between the difference of assessment metrics before and after alcohol consumption and BAC with |*r*| > 0.40

#### Linear Regression Models to Predict BAC

We used the metrics with |*r*| > 0.40 in Table 3 as input features for the linear regression models. We applied three linear regression models: least-squares regression, ridge regression, and LASSO regression to fit the training dataset, and evaluate their performance with Root Mean Square Error (RMSE). Their performance results are summarized in Table 4. The best model was the least-squares regression with a RMSE of 0.027. For reference, the mean BAC level measured at assessment, including before the drinking period, was 0.051 with a standard deviation of 0.057. We calculated a baseline RMSE, using the mean BAC as predicted values, which was 0.061. Our models demonstrate much better performance in predicting the actual BAC levels than the baseline method. For both the ridge regression and LASSO regression the features with the highest weights are the median of incongruent Stroop reaction time and median of digit symbol duration. However, for the least-squares regression model, the features with highest weights are the mean of digits correct per second and median of digit symbol duration. The weights of the input parameters for these models are shown in Figure 3.

**Table 4.** Performance of Linear, Ridge and LASSO models in predicting BAC. The baseline RMSE was calculated by using the mean BAC as every predicted value.

| Model | RMSE | L2 Penalty | L1 Penalty |
| --- | --- | --- | --- |
| Mean (baseline) | 0.061301 | -- | -- |
| Least-Square | 0.026769 | -- | -- |
| Ridge | 0.030692 | 10 | -- |
| LASSO | 0.029526 | -- | 0.001 |

**Figure 3.**
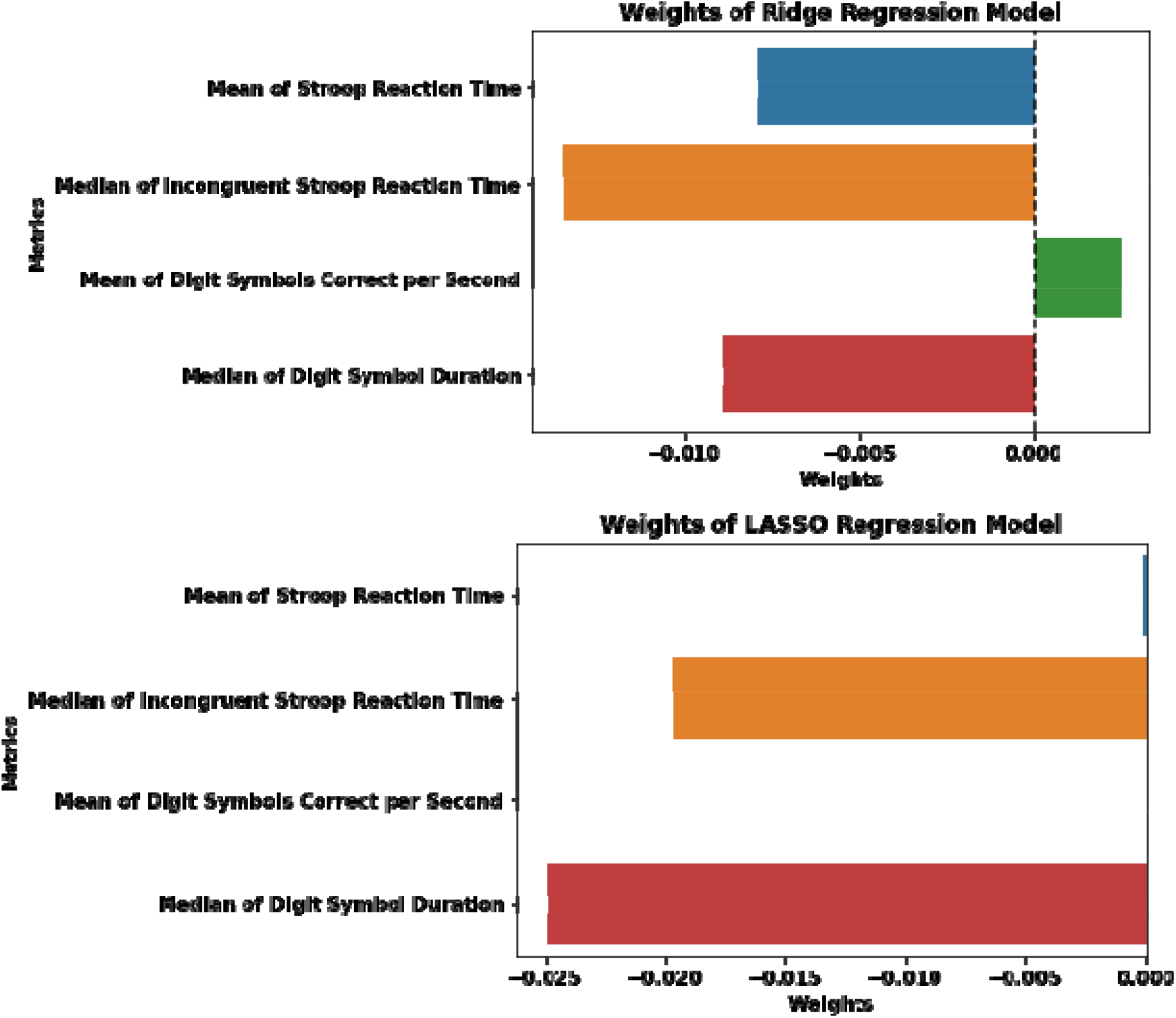
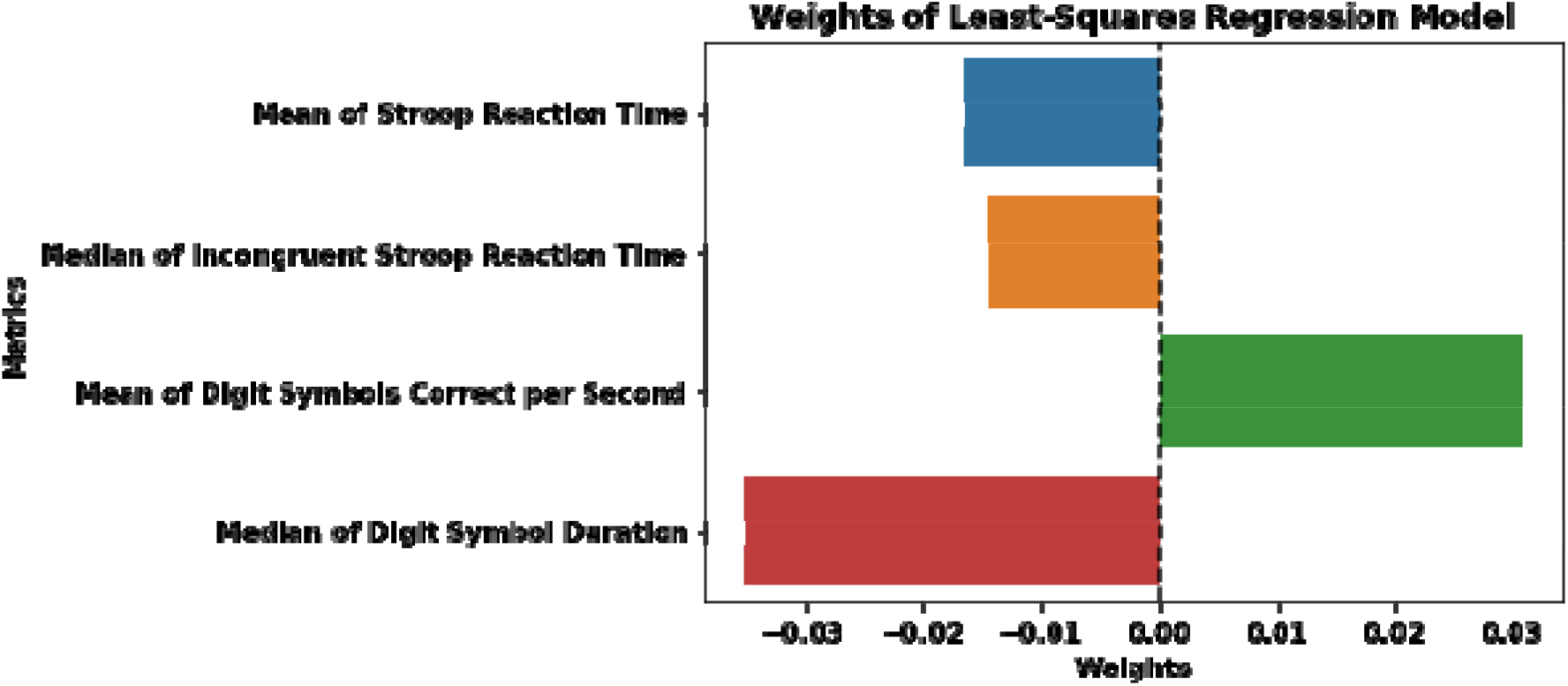
Weights of features for ridge, LASSO, and least-squares regression models

#### Logistic Regression Model for Classification

We used metrics with the lowest *p*-value from each assessment type to build a logistic regression model with regularizations to predict whether participants were either sober or intoxicated. These metrics were the mean of balance distance from center, mean of Stroop reaction time, median of Trails A duration time, mean of correct flanker reaction time, and median of digit symbol duration. All participant scores were inputted into this model. We reported the recall, precision, and accuracy for different penalties in Table 5. We found that the logistic regression model performed the best with an L1 penalty. This model had an accuracy of 80.65 % and a precision of 0.91. Additionally, this model has nonzero weights for mean of Trails A duration, median of Trails A duration, and the mean of correct flanker time reaction, shown in Figure 4. We also calculated a ROC curve for this model on the test dataset. As shown in Figure 5, the optimal performance for this model was at a threshold of 0.46 with a sensitivity of 73.3 %, a specificity of 75.0 %, and an area under the curve (AUC) of 0.86. In other words, if the model predicted probability for a subject is greater than 0.46, the subject is categorized to the intoxicated group.

**Table 5.** Performance of the logistic regression models.

| Penalty | Recall | Precision | Accuracy (%) |
| --- | --- | --- | --- |
| L2 | 0.47 | 0.78 | 67.74 |
| Elastic Net | 0.80 | 0.50 | 51.61 |
| L1 | 0.67 | 0.91 | 80.65 |

**Figure 4.**
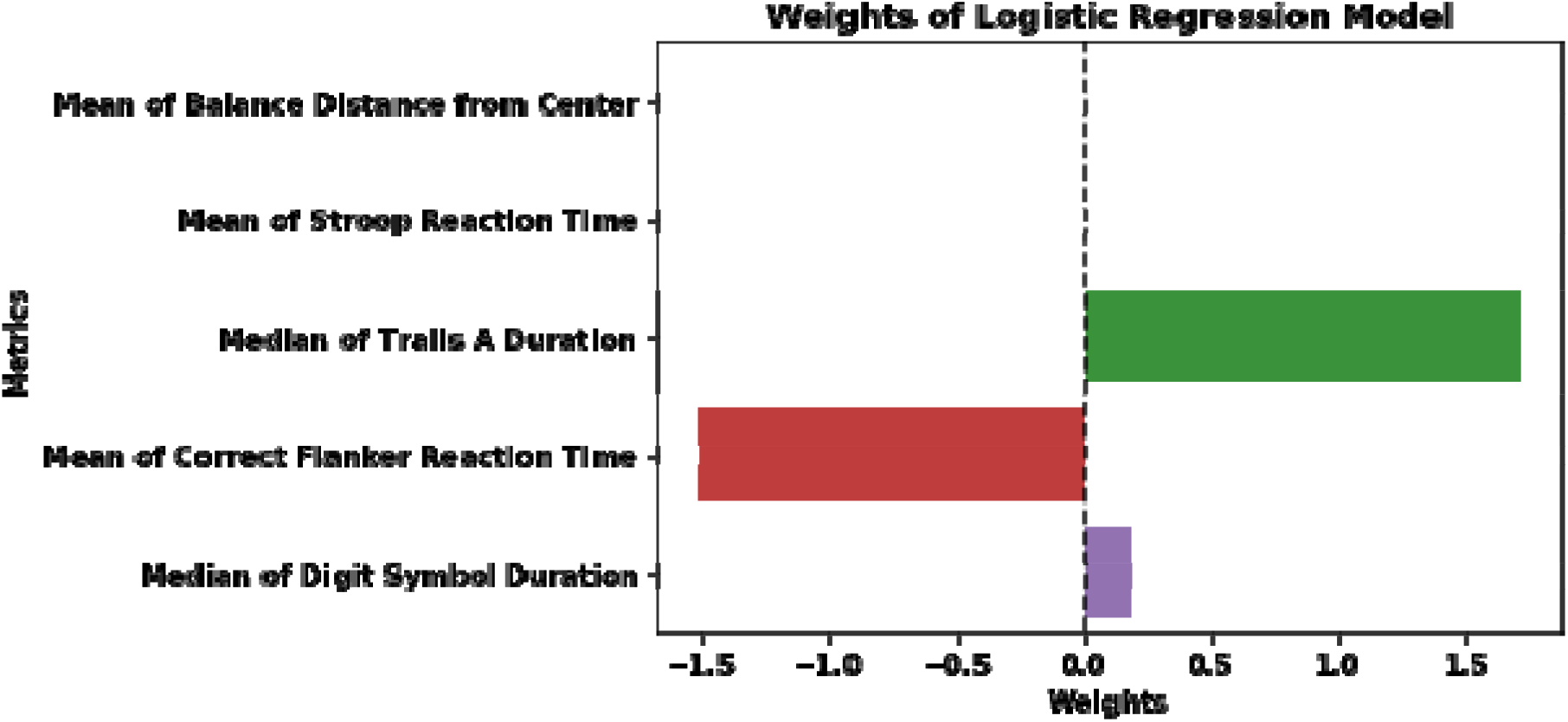
Weights of features for Logistic Regression Model with L1 Penalty

**Figure 5.**
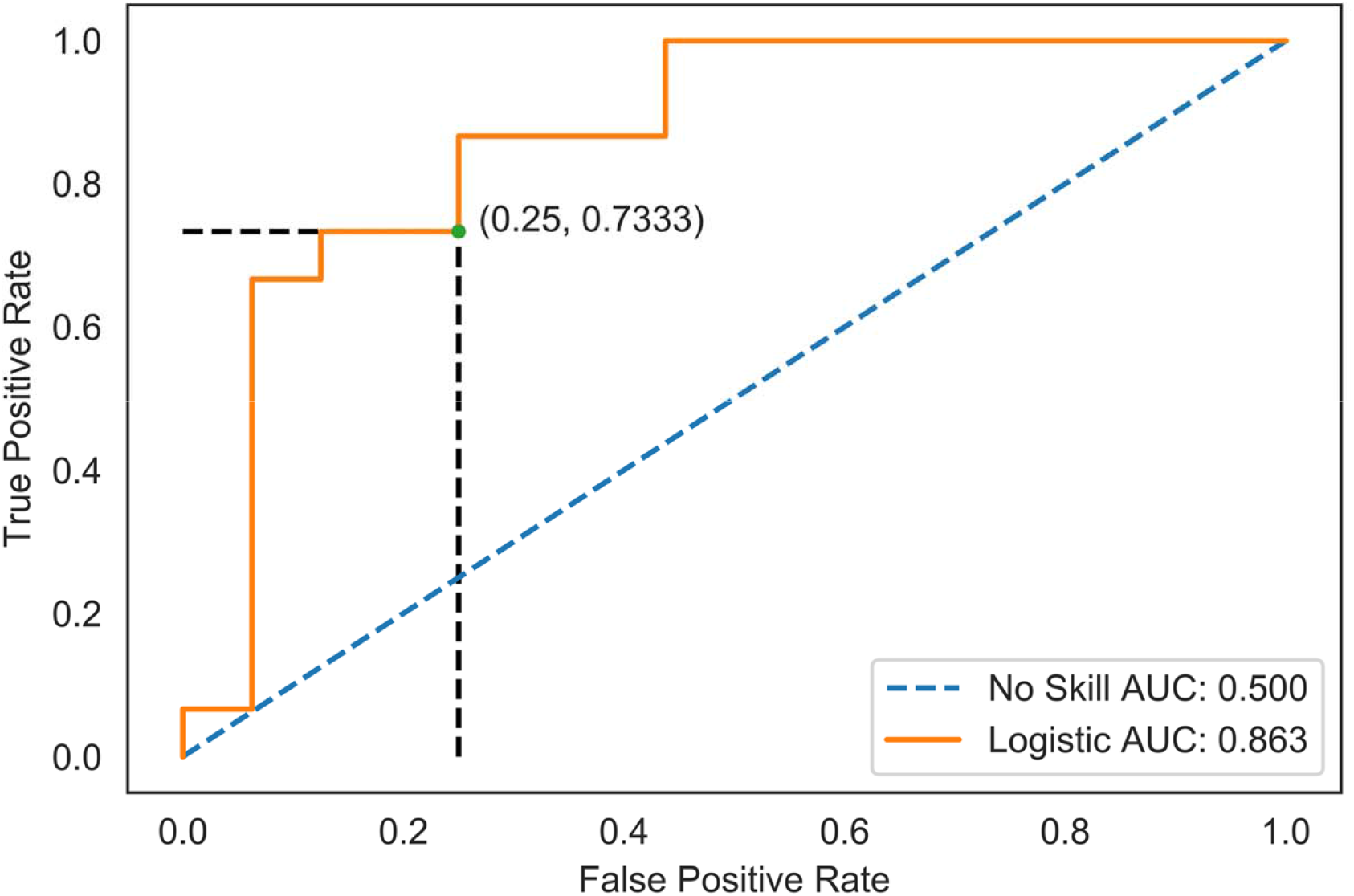
ROC Curve for logistic regression model with L1 penalty had an AUC OF 0.863. Optimal performance for this model was at a threshold of 0.46 marked by the black-dashed line with a sensitivity of 73.3 % and specificity of 75.0 %.

## Discussion

The findings of this study demonstrated that the BrainCheck battery has acceptable levels of accuracy in predicting BAC and in classifying sober and intoxicated participants. The RMSE for our linear regression model was 0.027 which is significantly lower than the baseline RMSE of 0.061. In classifying intoxicated vs. sober participants, our best logistic regression model performed at an accuracy of 80.65%, with good sensitivity (73.3%) and specificity (75.0%). These results showed the potential of the BrainCheck battery to detect cognitive impairment under alcohol consumption, in a similar way to use the BrainCheck sport battery in detecting mTBI (Yang et al., 2017).

Our results also demonstrated the moderate correlations between the changes in cognitive performance with the consumption of alcohol. The correlation between performance changes on the digit symbol substitution assessment and BAC levels is consistent with prior work that has shown impaired performance on the digit symbol substitution test with acute consumption of alcohol (Thapar et al., 1995). Additionally, the correlations observed between reaction time in the Stroop assessment and BAC levels are also consistent with impaired performance in previous studies (David et al., 2011). However, while we found significant differences in performance on Trails A, the previous work has observed only significant deficits in performance on Trails B, which is a more complex task than Trails A (Day et al., 2013). This different observation may be because we looked at an older population compared to the previous study (18-20 years old). The visuomotor performance defect by acute alcohol intoxication has been reported for older drinkers (Brumback et al., 2007).

There are further limitations to discuss. First, the participants represent a convenience sample and the number of alcoholic drinks consumed were not standardized/consistent across all the participants, due to avoiding the encouragement of alcohol consumption purely for the study. Also, some assessments may not be sensitive enough for changes in mild alcohol intoxication. Average BAC in this study was relatively low, which may not be sufficient to cause cognitive impairment for most individuals. In addition, assessments were administered on the same day and used the individuals’ baseline assessment as normal control, so we did not account for test-retest learning effects on participant assessment performance. Previous work on neuropsychological assessments demonstrate that participants generally perform better on the second assessment for both traditional and computerized tests (Cole et al., 2013; Hammers et al., 2011). This learning effect could mitigate the declined performance caused by alcohol, which increases the challenge to distinguish the alcohol usage based on the battery performance.

Compared to other methods of assessing an individual’s level of alcohol impairment, BrainCheck provides another option to assess level of alcohol impairment, including potential in predicting BAC and classifying intoxicated vs. sober. BrainCheck provides a shorter, gamified and portable test battery that can assess changes in cognitive function due to alcohol consumption, with the utility of being readily available on one’s smartphone or tablet.

## Data Availability

Data may be made available by contacting the corresponding author and with a data use agreement.

**Abbreviations**
BAC: blood alcohol content
mTBI: mild traumatic brain injury
*r*: Pearson correlation coefficient
LASSO: least absolute shrinkage selection operator
RMSE: root mean square error
ROC: receiver operator characteristic
AUC: area under the curve

## Acknowledgements

Study funding was provided by BrainCheck Inc. SJ would like to thank the University of Washington Undergraduate Program in Neural Computation and Engineering for funding his work in preparing the manuscript. HP would like to thank the Mary Gates Endowment for providing a research scholarship for his work in preparing the manuscript.

## Ethical Approval

This study protocol was reviewed and approved by Solutions IRB. All participants were required to sign for informed consent to be in the study.

## Data Availability

Data may be made available by contacting the corresponding author and with a data use agreement.

## Author Contributions

DME and YK contributed conception and design of the study; Data acquisition was performed by BF; SJ performed the statistical analysis, under the supervision of BH and RHG. HP wrote the first draft of the manuscript with assistance from SJ, which was revised and approved by BH and RHG.

## References

Bell, D. R., Guskiewicz, K. M., Clark, M. A., & Padua, D. A. (2011). Systematic Review of the Balance Error Scoring System. Sports Health, 3(3), 287–295. https://doi.org/10.1177/1941738111403122

Blakesley, R. E., Mazumdar, S., Dew, M. A., Houck, P. R., Tang, G., Reynolds, C. F., & Butters, M. A. (2009). Comparisons of Methods for Multiple Hypothesis Testing in Neuropsychological Research. Neuropsychology, 23(2), 255–264. https://doi.org/10.1037/a0012850

Bowie, C. R., & Harvey, P. D. (2006). Administration and interpretation of the Trail Making Test. Nature Protocols, 1(5), 2277–2281. https://doi.org/10.1038/nprot.2006.390

Brumback, T., Cao, D., & King, A. (2007). Effects of Alcohol on Psychomotor Performance and Perceived Impairment in Heavy Binge Social Drinkers. Drug and Alcohol Dependence, 91(1), 10–17. https://doi.org/10.1016/j.drugalcdep.2007.04.013

Cole, W. R., Arrieux, J. P., Schwab, K., Ivins, B. J., Qashu, F. M., & Lewis, S. C. (2013). Test–Retest Reliability of Four Computerized Neurocognitive Assessment Tools in an Active Duty Military Population. Archives of Clinical Neuropsychology, 28(7), 732–742. http s://doi.org/10.1093/arclin/act040

Collie, A., Maruff, P., Makdissi, M., McCrory, P., Bennell, K., & Darby, D. (2002). Quantifying the Cognitive Impairment Associated with Concussion: Using Blood Alcohol Concentration as a Reference Point. Medicine & Science in Sports & Exercise, 34(5), S94.

David, I. A., Volchan, E., Alfradique, I., Oliveira, L. de Pereira, M. G., Ranvaud, R., Vila, J., & Machado-Pinheiro, W. (2011). Dynamics of a Stroop matching task: Effect of alcohol and reversal with training. Psychology & Neuroscience, 4(2), 279–283. https://doi.org/10.3922/j.psns.2011.2.013

Day, A. M., Celio, M. A., Lisman, S. A., Johansen, G. E., & Spear, L. P. (2013). Acute and Chronic Effects of Alcohol on Trail Making Test Performance Among Underage Drinkers in a Field Setting. Journal of Studies on Alcohol and Drugs, 74(4), 635–641.

Eriksen, B. A., & Eriksen, C. W. (1974). Effects of noise letters upon the identification of a target letter in a nonsearch task. Perception & Psychophysics, 16(1), 143–149. https://doi.org/10.3758/BF03203267

Grant, S. A., Millar, K., & Kenny, G. N. C. (2000). Blood alcohol concentration and psychomotor effects. British Journal of Anaesthesia, 85(3), 401–406. https://doi.org/10.1093/bja/85.3.401

Groppell, S., Soto-Ruiz, K. M., Flores, B., Dawkins, W., Smith, I., Eagleman, D. M., & Katz, Y. (2019). A Rapid, Mobile Neurocognitive Screening Test to Aid in Identifying Cognitive Impairment and Dementia (BrainCheck): Cohort Study. JMIR Aging, 2(1). https://doi.org/10.2196/12615

Hammers, D., Spurgeon, E., Ryan, K., Persad, C., Heidebrink, J., Barbas, N., Albin, R., Frey, K., Darby, D., & Giordani, B. (2011). Reliability of Repeated Cognitive Assessment of Dementia Using a Brief Computerized Battery. American Journal of Alzheimer’s Disease & Other Dementiasr, 26(4), 326–333. https://doi.org/10.1177/1533317511411907

Hastie, T., Hastie, T., Tibshirani, R., & Friedman, J. H. (2001). The elements of statistical learning: Data mining inference, and prediction. New York: Springer.

Jaeger, J. (2018). Digit Symbol Substitution Test. Journal of Clinical Psychopharmacology, 38(5), 513–519. https://doi.org/10.1097/JCP.0000000000000941

NHTSA. (2019, December). 2018 Traffic Safety Facts: Alcohol Impaired Driving. https://crashstats.nhtsa.dot.gov/Api/Public/ViewPublication/812864

Riordan, B. C., Scarf, D., Moradi, S., Flett, J. A. M., Carey, K. B., & Conner, T. S. (2017). The accuracy and promise of personal breathalysers for research: Steps toward a cost-effective reliable measure of alcohol intoxication? Digital Health, 3. https://doi.org/10.1177/2055207617746752

Scarpina, F., & Tagini, S. (2017). The Stroop Color and Word Test. Frontiers in Psychology, 8 https://doi.org/10.3389/fpsyg.2017.00557

Schober, P., Boer, C., & Schwarte, L. A. (2018). Correlation Coefficients: Appropriate Use and Interpretation. Anesthesia & Analgesia, 126(5), 1763–1768. https://doi.org/10.1213/ANE.0000000000002864

Schweizer, T. A., Vogel-Sprott, M., Danckert, J., Roy, E. A., Skakum, A., & Broderick, C. E. (2006). Neuropsychological Profile of Acute Alcohol Intoxication during Ascending and Descending Blood Alcohol Concentrations. Neuropsychopharmacology, 31(6), 1301–1309. https://doi.org/10.1038/sj.npp.1300941

Stuster, J. (1998). Validation of the Standardized Field Sobriety Test Battery at BACs Below 0.10 Percent: Final Report. National Highway Traffic Safety Administration.

Thapar, P., Zacny, J. P., Thompson, W., & Apfelbaum, J. L. (1995). Using Alcohol as a Standard to Assess the Degree of Impairment Induced by Sedative and Analgesic Drugs Used in Ambulatory Surgery. Anesthesiology: The Journal of the American Society of Anesthesiologists, 82(1), 53–59.

Valenzuela, C. F. (1997). Alcohol and Neurotransmitter Interactions. RESEARCH WORLD, 21(2), 5.

Yang, S., Flores, B., Magal, R., Harris, K., Gross, J., Ewbank, A., Davenport, S., Ormachea, P., Nasser, W., Le, W., Peacock, W. F., Katz, Y., & Eagleman, D. M. (2017). Diagnostic accuracy of tablet-based software for the detection of concussion. PLoS ONE, 12(7). https://doi.org/10.1371/journal.pone.0179352

